# Sarcopenia, adiposity, and discrepancies in cystatin C versus creatinine-based eGFR in patients with cancer: a cross-sectional study

**DOI:** 10.1101/2023.02.08.23285587

**Authors:** Paul E. Hanna, Tianqi Ouyang, Ismail Tahir, Nurit Katz-Agranov, Qiyu Wang, Lea Mantz, Ian Strohbehn, Daiana Moreno, Destiny Harden, James E. Dinulos, Duru Cosar, Harish Seethapathy, Justin F. Gainor, Sachin J. Shah, Shruti Gupta, David E. Leaf, Florian J. Fintelman, Meghan E. Sise

## Abstract

**Purpose:** Creatinine-based estimated glomerular filtration rate (eGFR_CRE_) may overestimate kidney function in patients with sarcopenia. While Cystatin C-based eGFR (eGFR_CYS_) is less affected by muscle mass, it may underestimate kidney function in patients with obesity. We sought to evaluate the relationship between body composition and discrepancies between creatinine and eGFR_CRE_ and eGFR_CYS_ in patients with cancer.

**Methods:** We conducted a cross-sectional study of consecutive adults with cancer who had an abdominal CT scan performed within 90 days of simultaneous eGFR_CRE_ and eGFR_CYS_ measurements between May 2010-January 2022. Sarcopenia was defined using independent sex-specific cutoffs for skeletal muscle index (SMI) at the level of the third lumbar vertebral body (<39 cm^2^/m^2^ for women,<55 cm^2^/m^2^ for men). High adiposity was defined as the highest sex-specific quartile of total (visceral plus subcutaneous) adiposity index in the cohort. The primary outcome was eGFR discrepancy, defined by eGFR_CYS_ >30% lower than eGFR_CRE_. We estimated the odds of eGFR discrepancy using multivariable logistic regression modeling.

**Results:** Of 545 included patients (mean age 63 ±14 years, 300 [55%] females, 440 [80.7%] non-Hispanic white), 320 (58.7%) met the criteria for sarcopenia and 136 (25%) had high adiposity. After adjustment for potential confounders, sarcopenia and high adiposity were both associated with >30% eGFR discrepancy (adjusted odds ratio [aOR] 1.90, 95% confidence interval [CI] 1.12–3.24; aOR 2.01, 95% CI 1.15–3.52, respectively).

**Conclusion:** Discrepancies in eGFR_CRE_ and eGFR_CYS_ are common in adult patients with cancer, and sarcopenia and high adiposity are both independently associated with large eGFR discrepancies.

**Significance statement:** Serum creatinine may overestimate glomerular filtration rate (GFR) in patients with muscle loss, which is particularly common among patients with cancer. Serum cystatin C may perform better than creatinine in such patients, but its accuracy is affected by obesity. We performed body composition analysis using computed tomography scans in 545 adult patients with cancer and found that both sarcopenia and high adiposity were independently associated with greater discrepancies in serum creatinine-vs. cystatin C-based estimated GFR. These findings highlight the need for future studies to improve and personalize GFR assessment in patients with cancer, particularly in those who will receive renally cleared medications and anti-neoplastic therapies with a narrow therapeutic index.

## Introduction

Accurate assessment of estimated glomerular filtration rate (eGFR) is important, especially in patients with cancer who commonly receive renally-cleared medications with narrow therapeutic indices. Estimating GFR using serum creatinine (SCr) remains the most widely used method in both clinical practice and research.^1-3^ Yet, because creatinine is a byproduct of muscle metabolism, it can overestimate kidney function in patients with sarcopenia, a common condition in patients with cancer.^4,5^ Cystatin C is a low molecular weight (13K Dalton) protein produced by all nucleated cells that is filtered by the glomerulus and is not reabsorbed or secreted.^6^ Unlike SCr, cystatin C is not readily affected by age, sex, muscle mass, or diet, and has been increasingly used as an alternative to SCr to estimate GFR, with some caveats (e.g., cystatin C is elevated in obesity and acute inflammatory states).^1,4,7,8^

Cancer cachexia is a multifactorial syndrome characterized by loss of skeletal muscle leading to progressive functional impairment, which is strongly associated with increased mortality.^9^ However, recent studies have shown that visibly wasted patients are increasingly rare in clinical practice; a clinical phenotype of “sarcopenic obesity” has emerged to describe patients with high body mass index (BMI) but with underlying depleted muscle mass.^10,11^ Therefore, clinical impression of cachexia or BMI are inadequate markers for sarcopenia in patients with cancer, and more precise evaluation of body composition using imaging techniques has been increasingly used in clinical practice over the past 15 years.^12,13^ Computed tomography (CT) scans are considered a gold standard for body composition assessment;^14,15^ the cross-sectional areas of tissues in single images at the level of the third lumbar vertebra (L3) strongly correlate with whole body adipose tissue, muscle, and lean tissue mass (**Figure 1**).^16,17^ Using different diagnostic thresholds, studies have shown that 40 to 60% of patients with locally advanced or metastatic cancer had imaging-defined sarcopenia.^18-22^ In addition, imaging-defined sarcopenia has been shown to have a strong association with adverse clinical outcomes in patients with cancer.^23-25^

**Figure 1.**
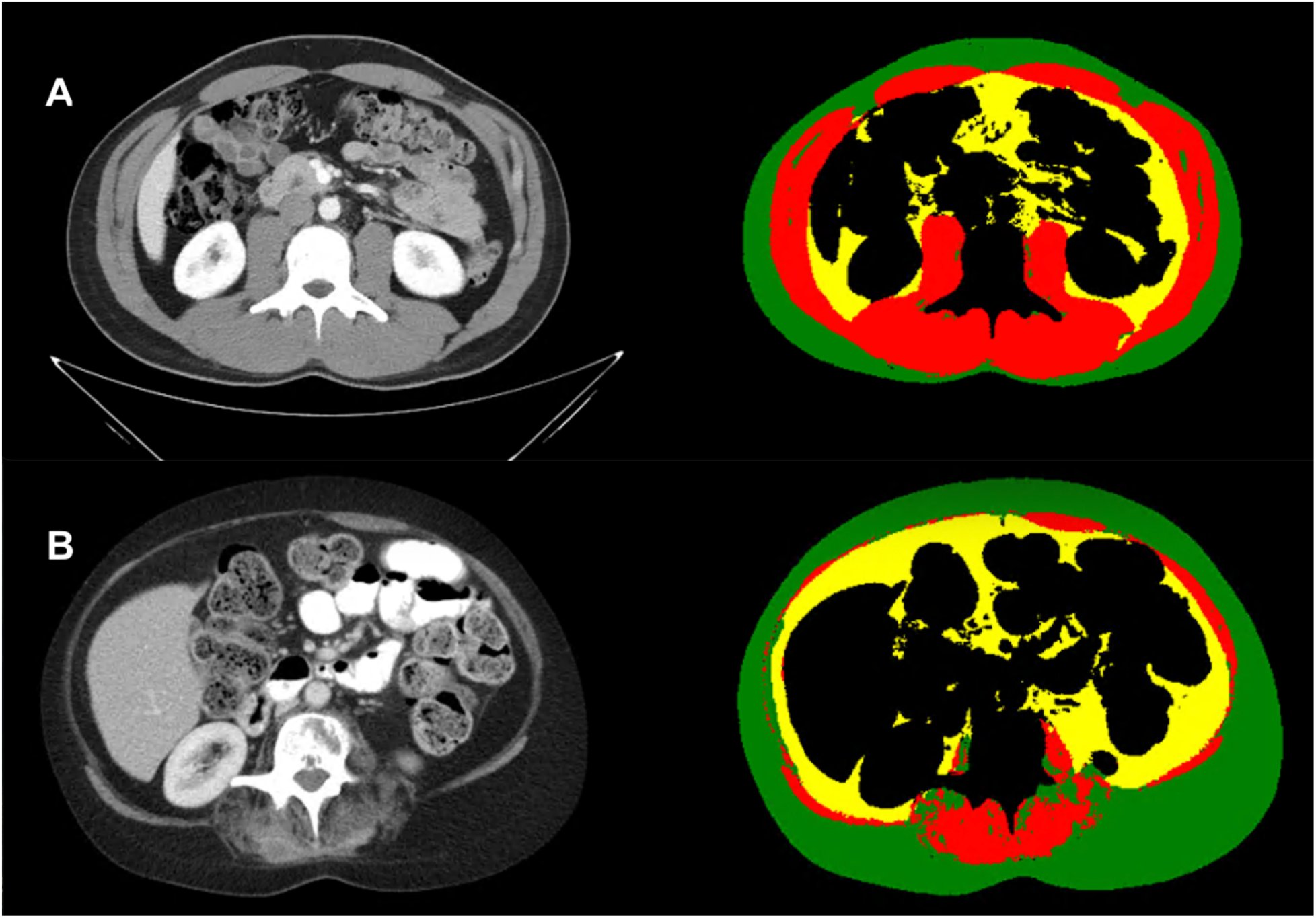
Body composition analysis at the level of the third lumbar vertebral body. Representative axial computed tomography images at the level of the 3^rd^ lumbar vertebral body (left column) and matching segmentation label maps (right column). Patient A was a 26 to 30-year-old male with a skeletal muscle [red] index (SMI) of 65.9 (4^th^ quartile), subcutaneous adiposity [green] index (SAI) of 38.6 (2^nd^ quartile) and visceral adiposity [yellow] index (VAI) of 25.8 (1^st^ quartile). Patient B is a 46 to 50-year-old female with SMI [red] of 18.9 (1^st^ quartile), SAI [green] of 77.3 (2^nd^ quartile) and VAI [yellow] of 36.92 (3^rd^ quartile).

Given that patients with active cancer undergo frequent CT scans for diagnosis and monitoring of disease progression, CT is a convenient tool to evaluate body composition and detect sarcopenia, and may aid in the accurate interpretation of SCr or cystatin C-based eGFR. Because patients with sarcopenia may have a falsely-low SCr and obesity is associated with an elevated cystatin C, we hypothesized that imaging-defined sarcopenia and high adiposity index are both associated with large discrepancies between SCr and cystatin C-based eGFR (eGFR_CRE_ and eGFR_CYS_). Our objective was to evaluate the association between body composition (skeletal muscle index [SMI] and adiposity) assessed by abdominal CT scan and discrepancies between SCr and cystatin C-based eGFR (eGFR_CRE_ and eGFR_CYS_) in patients with cancer.

## Methods

### Patient population and study design

Using Mass General Brigham’s centralized data warehouse, the Research Patient Data Registry (RPDR),^26,27^ we designed a cross-sectional study of consecutive adult patients with a pre-existing diagnosis of malignancy who had simultaneous measurement of SCr and cystatin C as a part of routine care between May 2010 and January 2022. We defined the baseline as the date of the simultaneous eGFR_CRE_ and eGFR_CYS_ measurement. In patients with multiple simultaneous assessments, the first instance was used. We calculated eGFR_CRE_ using the CKD Epidemiology Collaboration (CKD-EPI) 2021 race-free equation,^4^ while eGFR_CYS_ was calculated using the CKD Epidemiology Collaboration (CKD-EPI) 2012 race-free equation.^28,29^ We included all patients who had an abdominal computed tomography (CT) scan performed within 90 days before or after the baseline date. We excluded patients with eGFR_CRE_ < 15mL/min/1.73m^2^ and those whose CT scans failed quality control for body composition analysis (**Supplemental Table 1**).

**Table 1.**
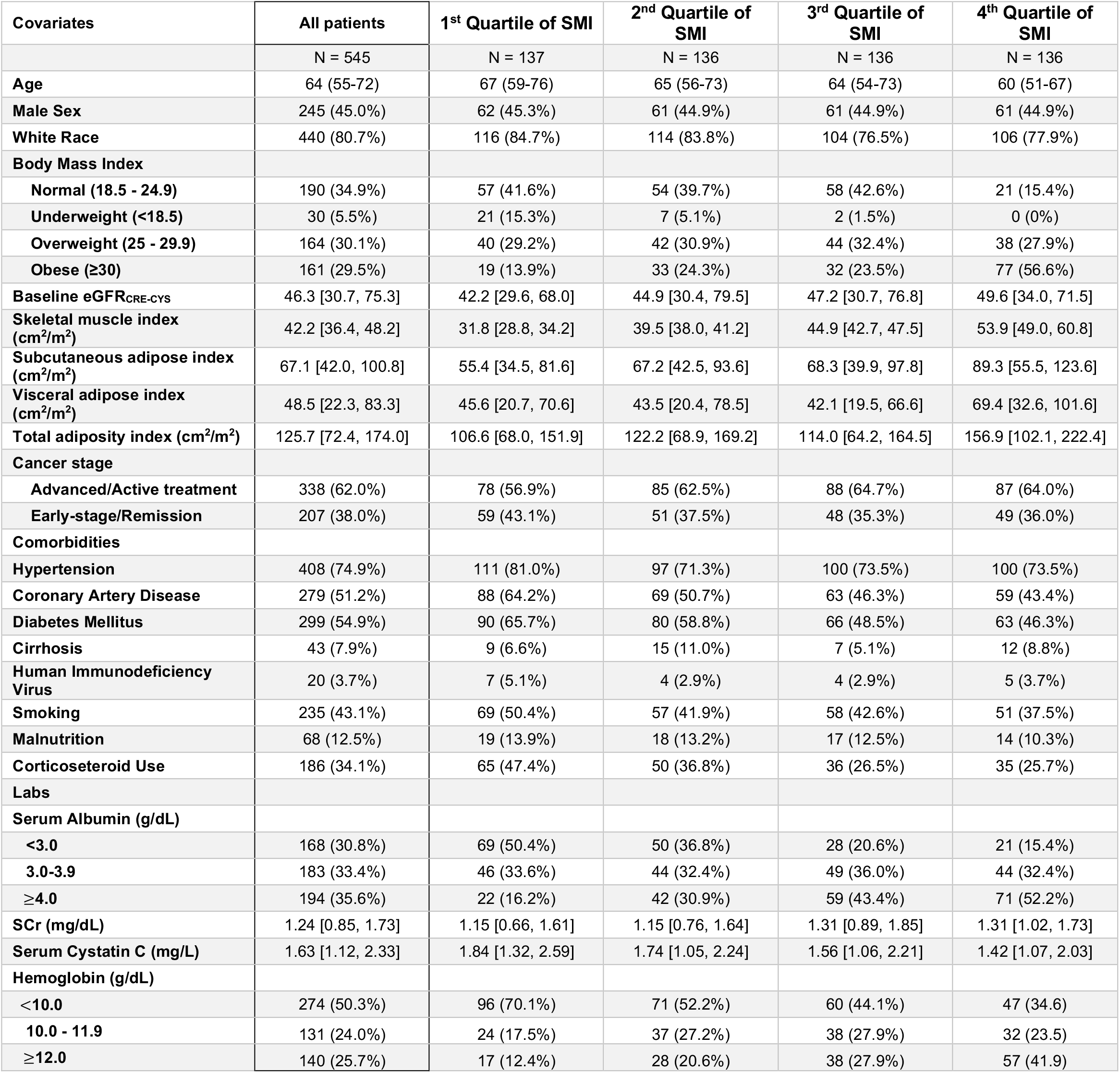
Baseline patients characteristics. Count and percent or median interquartile ranges are shown. Abbreviations: eGFR = estimated glomerular filtration rate, ACEi/ARB = Angiotensin Converting Enzyme Inhibitor/Angiotensin Receptor Blocker. Missing data: Serum albumin is missing for 4 patients (0.7%). Visceral adipose index is missing for 9 patients (1.7%) and subcutaneous adipose index is missing for 77 patients (14.1%)

### Data collection

Comorbidities were defined based on diagnosis codes appearing any time prior to the baseline date. Corticosteroid use was defined by active prescription within 30 days of the baseline date. Cancer type was determined by the most frequently coded cancer-related diagnosis prior to the baseline. Cancer stage was determined by chart review and defined as early-stage vs. locally advanced or advanced (stage 3 or 4) based on oncologists’ documentation. Patients who had previously completed antineoplastic treatment or underwent surgery without evidence of disease as documented by their oncologist were considered to be in remission. Baseline chronic kidney disease was defined by the 2021 race free CKD-EPI equation that incorporates both SCr and cystatin C.^7^

### Body composition analysis on CT scans

We used a multistep pipeline including previously validated machine learning algorithms to identify the third lumbar vertebral level (L3) on axial images and segment the cross-sectional area of skeletal muscle and adipose tissue at this level.^12,30-33^ Each segmentation label map was independently reviewed by a trained analyst with two years’ experience performing body composition analysis blinded to clinical outcomes including eGFR_CRE_ and eGFR_CYS_ (I.T.). We calculated SMI (cm^2^/m^2^) by dividing the skeletal muscle cross sectional area by the patient’s height squared. Visceral and subcutaneous fat areas were normalized for the height (divided by height squared) to calculate visceral adiposity index (VAI, cm^2^/m^2^) and subcutaneous adiposity index (SAI, cm^2^/m^2^) (**Figure 1**). Total adiposity index was calculated by adding VAI and SAI.^12,33^ When possible, we selected outpatient CT scans; if no outpatient CT scan was available within 90 days, we included the inpatient CT scan closest to the baseline date.

### Primary exposure

Sarcopenia was defined using previously published SMI cutoffs of less than 39 cm^2^/m^2^ for women and less than 55 cm^2^/m^2^ for men.^12^ High adiposity was defined as the highest sex-specific quartile of total adiposity index (VAI plus SAI), as no cutoffs for low or high adiposity index exist.

### Primary outcomes

The primary outcome was eGFR discrepancy, defined as eGFR_CYS_ more than 30% lower than the eGFR_CRE_. The reference group consisted of all patients who did not meet criteria for eGFR discrepancy. The reference group included those whose eGFR_CYS_ was more than 30% greater than eGFR_CRE_ since as discrepancy in this direction would not place patients at high risk of adverse outcomes related to eGFR overestimation.^34^ The 30% cut-off was chosen as it is commonly used in clinical studies to define the accuracy of eGFR from measured GFR.^35,36^

### Secondary outcomes

To evaluate a more severe eGFR discrepancy, we additionally identified a subset of patients with eGFR_CYS_ more than 50% lower than eGFR_CRE_. Additionally, to assess the absolute eGFR difference (eGFR_DIFF_), we subtracted eGFR_CRE_ from eGFR_CYS_; negative values indicated that the eGFR_CRE_ was higher than the eGFR_CYS_. We evaluated the continuous relationship between eGFR_DIFF_ and SMI.

### Statistical Analysis

We reported baseline characteristics for the entire cohort by SMI quartile using counts and percentages for categorical variables, means with standard deviations (±SD) for normally distributed continuous variables, and median and interquartile range (IQR) for skewed variables. Variables with missing values were imputated using multiple imputation method; five imputed data sets were obtained through chained equations, parameter estimates from each imputed dataset were pooled using Rubin’s rules.^37^

We examined the unadjusted associations between baseline demographics, comorbidities, medications, laboratory studies, and body composition (sarcopenia and high adiposity), with eGFR discrepancy. We fit a multivariable logistic regression model to determine the association between eGFR discrepancy and body composition accounting for potential confounders. Hemoglobin and serum albumin were evaluated in clinically relevant categories. Unadjusted spline regression was performed to evaluate the relationship between SMI and eGFR_DIFF_ (eGFR_CYS_ minus eGFR_CRE_) using B-Spline basis for cubic polynomial splines with three degrees of freedom.

To address bias, we also performed sensitivity analyses excluding patients with inpatient CT scans, liquid tumors(lymphoma, leukemia, or myeloma), and acute kidney injury (AKI) at the time of eGFR_CRE_ and eGFR_CYS_ assessment, and by. AKI was defined as a SCr that was 50% higher than the lowest SCr in the 365 days prior to baseline.

All comparisons were two-sided, with *p* < 0.05 considered significant. All analyses were performed using R 4.1.1 (R Foundation), SAS 9.4 (SAS Institute), and GraphPad PRISM V.9.1.0 (GraphPad Software). The design and reporting of this cross-sectional study followed the Strengthening the Reporting of Observational Studies in Epidemiology (STROBE) guidelines for reporting observational studies.

### Informed Consent

The Massachusetts General Brigham Institutional Review Board approved this study and waived the need for informed consent.

## Results

### Patient and CT scan characteristics

Of the 1,988 patients with cancer originally included in this study, 662 had one or more abdominal CT scans within 90 days of the baseline date. We excluded 46 patients with eGFR_CRE_ < 15mL/min/1.73m^2^ and 48 patients whose scans failed image segmentation or quality control (**Figure 2, Supplemental Table 1**). The included 545 patients had a median age of 64 years (interquartile range [IQR] 55 - 72), 45% were male, and 80.7% were non-Hispanic white (**Table 1**). Patients had a wide array of cancers types (**Supplemental Table 2**); the most common cancer types were breast (12.5%), gynecological (11.4%), and gastrointestinal (10.8%). Most patients (N = 338; 62%) had advanced disease. The mean BMI was 27.6 kg/m^2^ (SD ± 7.2). Most CT scans (N = 375; 69%) were performed in the outpatient setting and N = 334 (61%) were performed with intravenous contrast.

**Table 2.** Predictors of eGFR discrepancy. eGFR discrepancy is defined by eGFR_CYS_ >30% lower than eGFR_CRE_. Baseline eGFR_CRE-CYS_ was defined using the race-free CKD Epi 2021 combined cystatin C and creatinine equation. Abbreviations: CI = confidence interval, REF = reference, ACEi/ARB = angiotensin converting enzyme inhibitor or angiotensin II receptor blockade, HIV = human immunodeficiency virus.

| Covariates | 30% eGFR discrepancy<br>(N= 259) vs. reference (N=286) |  |  |  |  |  |
| --- | --- | --- | --- | --- | --- | --- |
|  | Unadjusted |  |  | Multivariable |  |  |
|  | OR | 95% CI | p-value | Adj OR | 95% CI | p-value |
| **Age (per 10 years)** | 1.15 | 1.02, 1.30 | 0.024 | 0.92 | 0.77, 1.10 | 0.344 |
| **Male sex** | 1.33 | 0.95, 1.87 | 0.1 | 0.69 | 0.41, 1.15 | 0.154 |
| **Baseline eGFR<sub>CRE-CYS</sub>** | 0.98 | 0.97, 0.98 | <0.001 | 0.98 | 0.97, 0.99 | <0.001 |
| **Sarcopenia** | 2.38 | 1.67, 3.38 | <0.001 | 1.90 | 1.12, 3.24 | 0.018 |
| **Highest adiposity quartile** | 1.32 | 0.89, 1.96 | 0.160 | 2.01 | 1.15, 3.52 | 0.014 |
| **Liquid vs. solid tumor** | 2.83 | 1.78, 4.48 | <0.001 | 1.51 | 0.80, 2.84 | 0.201 |
| **Acute kidney injury** | 1.93 | 1.37, 2.73 | <0.001 | 0.56 | 0.34, 0.92 | 0.022 |
| **Inpatient vs. Outpatient CT scan** | 4.60 | 3.09, 6.84 | <0.001 | 1.81 | 1.09, 3.00 | 0.021 |
| **Smoking** | 1.54 | 1.09, 2.17 | 0.014 | 1.11 | 0.70, 1.76 | 0.649 |
| **Comorbidities** |  |  |  |  |  |  |
| **Hypertension** | 2.57 | 1.70, 3.88 | <0.001 | 1.00 | 0.54, 1.87 | 0.988 |
| **Coronary Artery Disease** | 2.65 | 1.87, 3.75 | <0.001 | 1.08 | 0.67, 1.74 | 0.760 |
| **Diabetes Mellitus** | 4.03 | 2.81, 5.79 | <0.001 | 1.42 | 0.86, 2.33 | 0.170 |
| **Cirrhosis** | 4.65 | 2.18, 9.92 | <0.001 | 3.04 | 1.22, 7.59 | 0.017 |
| **HIV** | 1.69 | 0.68, 4.21 | 0.26 |  |  |  |
| **Malnutrition** | 1.68 | 1.00, 2.82 | 0.048 | 1.02 | 0.53, 1.96 | 0.957 |
| **Thyroid disease** | 1.52 | 1.04, 2.24 | 0.032 | 0.97 | 0.59, 1.61 | 0.920 |
| **Medication Use\*** |  |  |  |  |  |  |
| **Corticosteroids** | 3.57 | 2.45, 5.19 | <0.001 | 1.71 | 1.06, 2.76 | 0.029 |
| **Labs** |  |  |  |  |  |  |
| **Albumin (g/dL)** |  |  |  |  |  |  |
| **<3.0** | REF | — |  | — | — |  |
| **3.0-3.99** | 5.58 | 3.47, 8.99 | <0.001 | 3.28 | 1.79, 6.02 | <0.001 |
| **≥4.0** | 15.6 | 9.28, 26.1 | <0.001 | 9.00 | 4.46, 18.2 | <0.001 |
| **Hemoglobin (g/dL)** |  |  |  |  |  |  |
| **<10.0** | REF | — |  |  |  |  |
| **10.0 – 11.99** | 1.70 | 1.00, 2.89 | 0.052 | 0.73 | 0.36, 1.47 | 0.375 |
| **≥12.0** | 6.31 | 3.97, 10.0 | <0.001 | 1.22 | 0.61, 2.45 | 0.566 |

**Figure 2.**
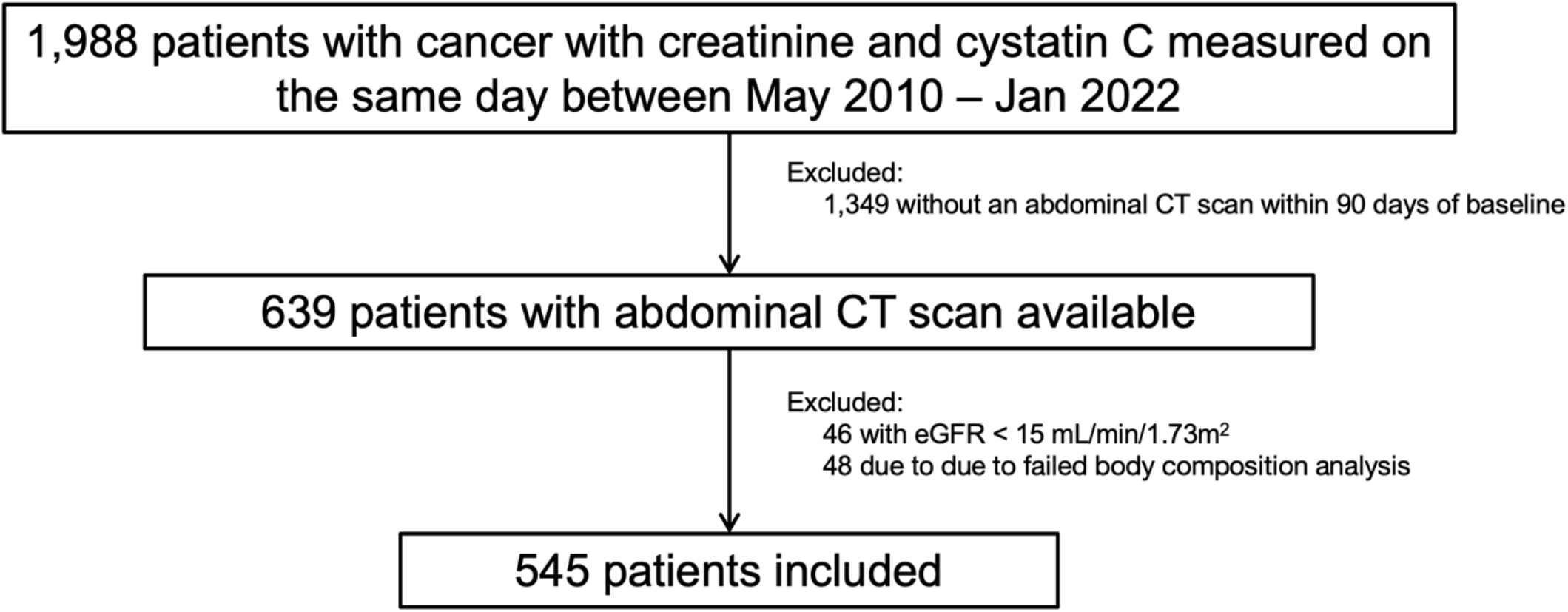
Patient flow. Abbreviations: CT = computed tomography, eGFR = estimated glomerular filtration rate.

### Body composition analysis

SMI was available for all included patients. A total of 320 (58.7%) patients met predefined sex-specific SMI thresholds for sarcopenia.^12^ Though mean SMI was higher in men compared to women (45.8 ± 11.7 cm^2^/m^2^ vs. 40.5 ± 8.6 cm^2^/m^2^, *p* < 0.001), a higher fraction of men met criteria for sarcopenia (81.5% vs. 40%; *p* < 0.001) (**Figure 3**). Patients that self-identified as Asian more commonly met imaging criteria for sarcopenia compared to other races/ethnicities (**Supplemental Figure 1**).

**Figure 3.**
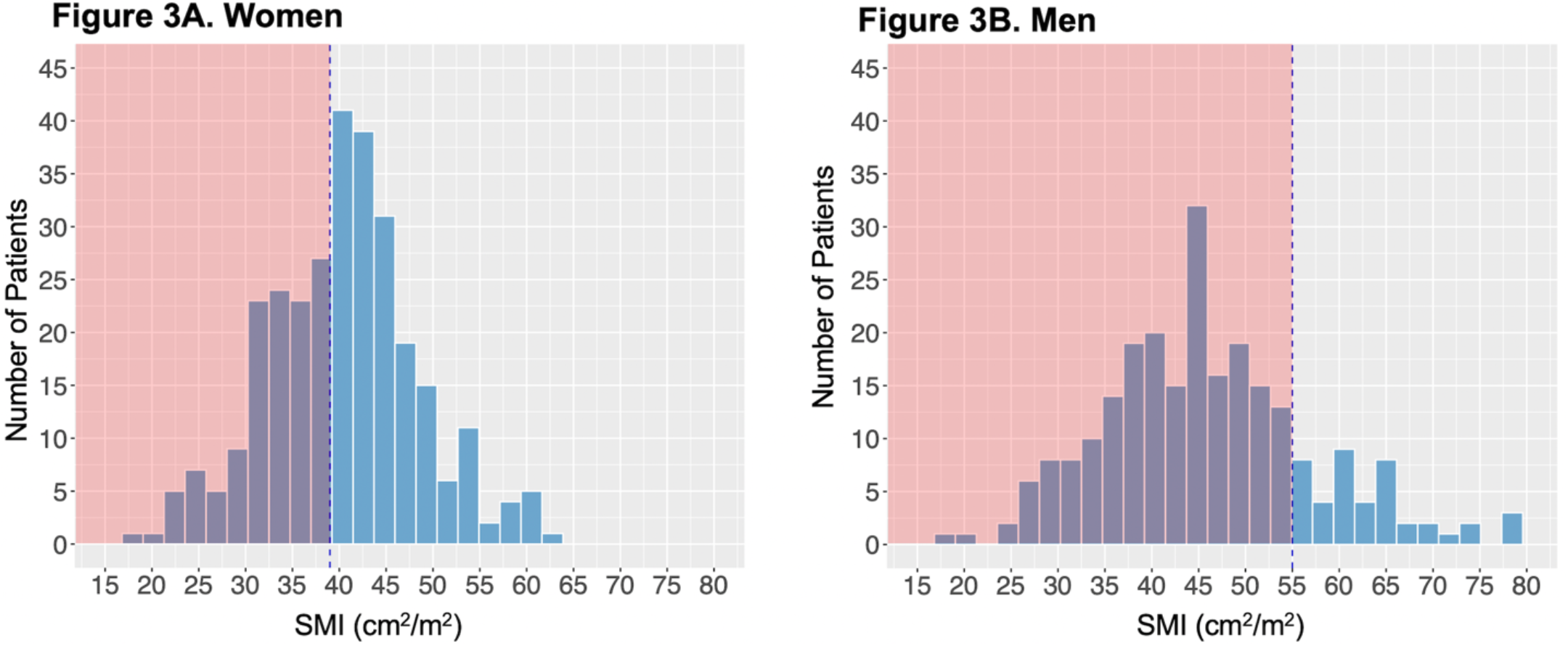
Histograms showing the distribution of skeletal muscle index by sex. The red shaded area denotes the sex-specific cutoffs for sarcopenia as described by Prado et al.^12^ Abbreviations SMI = skeletal muscle index

Visceral and subcutaneous fat indices were available for 536 (98%) and 468 (86%) of the cohort, respectively, and both were available in 465 (85%) of the cohort. Mean VAI was higher in men than women (mean 69.5± 43.0 cm^2^/m^2^ vs. 44.7 ± 35.8 cm^2^/m^2^; *p* < 0.001), whereas mean SAI was higher in women than men (mean 89.0 ± 54.7 cm^2^/m^2^ vs. 62.1 ± 40.1 cm^2^/m^2^; *p* < 0.001) (**Supplemental Figure 2**).

### Primary and secondary outcomes

A total of 259 patients (48%) had a >30% eGFR discrepancy. A scatterplot of eGFR_CRE_ vs. eGFR_CYS_ is shown in **Supplemental Figure 3A** and the distribution of the differences between eGFR_CRE_ and eGFR_CYS_ is shown in **Supplemental Figure 3B**. Among the 320 patients with sarcopenia, 180 (56.3%) met the primary outcome, among the 136 patients with high adiposity 72 (52.9%) met the primary outcome, and 41 (68.3%) of the 60 patients with both sarcopenia and high adiposity met the primary outcome (**Figure 4**). In the final multivariable model adjusted for variables shown in **Table 2**, sarcopenia and high adiposity each remained independent predictors of eGFR discrepancy (aOR 1.90, 95% CI 1.12 - 3.24; p = 0.018) and (aOR 2.01, 95% CI 1.15 - 3.52; p = 0.014), respectively (**Figure 5**).

**Figure 4.**
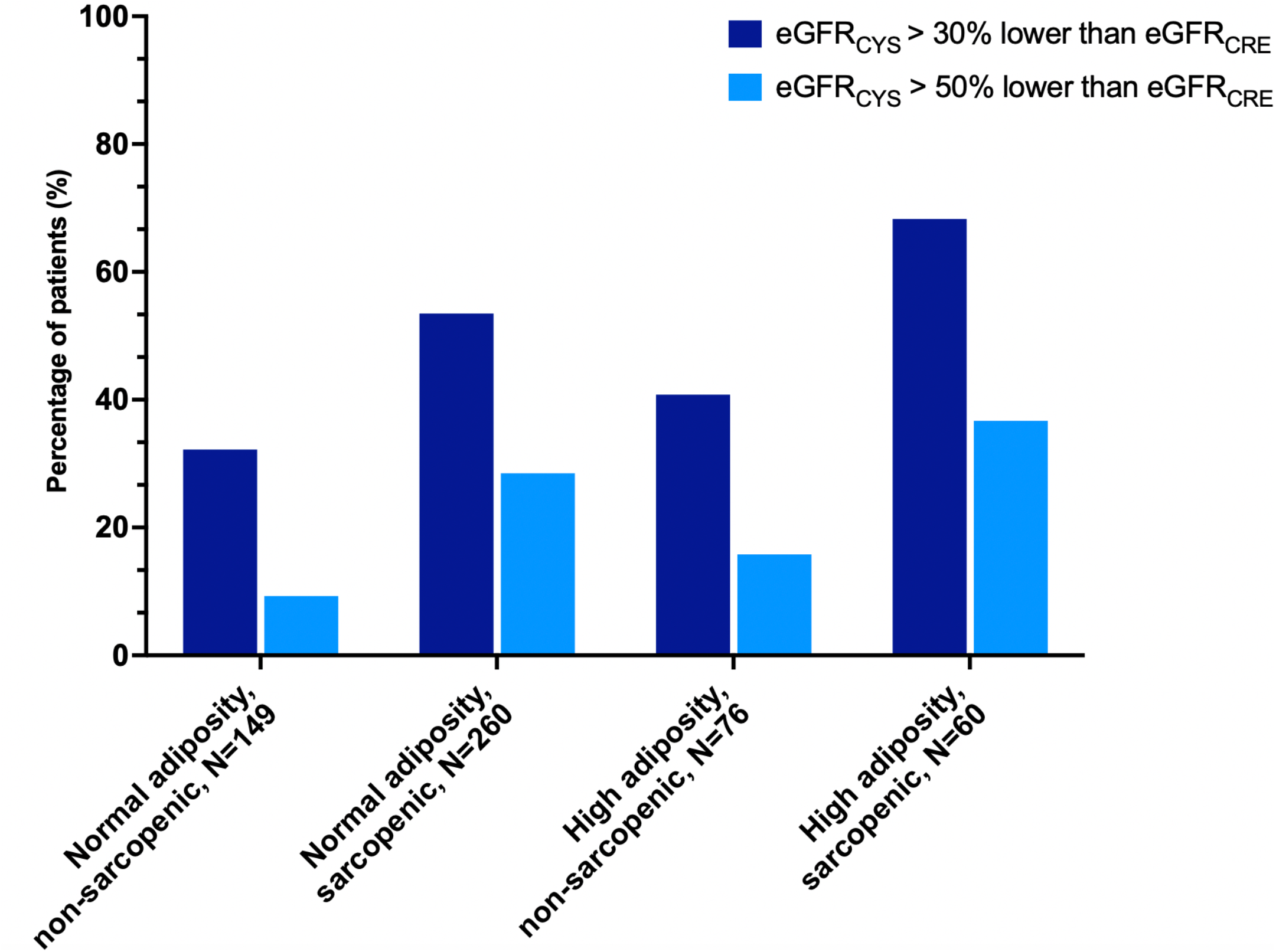
Rate of eGFR discrepancy by body composition. The rate of eGFR discrepancy (eGFR_CYS_ > 30% lower than eGFR_CRE_) and severe eGFR discrepancy (eGFR_CYS_ > 50% lower than eGFR_CRE_) by body composition. Sarcopenia is defined by sex-specific cutoffs for skeletal muscle index (<39cm^2^/m^2^ for women and <55cm^2^/m^2^ for men); high adiposity is defined by the highest sex-specific quartile.

**Figure 5.**
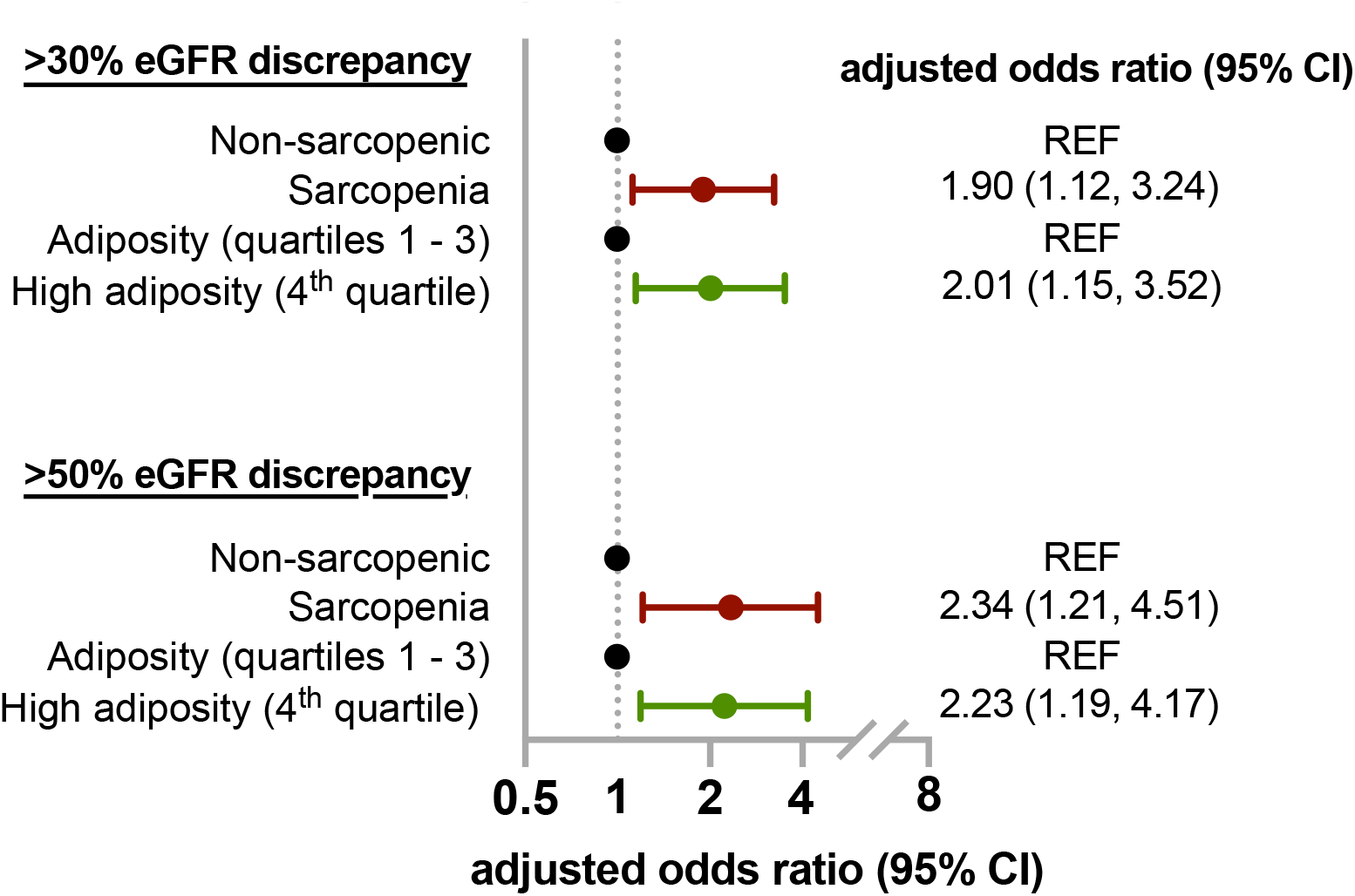
Forrest plot illustrating the strength of associations with 30% and 50% eGFR discrepancies. The final model for both >30% and >50% eGFR discrepancy are adjusted the variables shown in **Table 2**. The unadjusted and multivariable logistic regression model for >50% eGFR discrepancy is shown in **Supplemental Table 4**. Abbreviations: eGFR = estimated glomerular filtration rate, REF = reference group, CI = confidence interval

Sarcopenia and high adiposity were also strongly associated with having an eGFR_CYS_ more than 50% lower than eGFR_CRE_ (aOR 2.34, 95% CI 1.21 – 4.51; p = 0.012 and aOR 2.23, 95% CI 1.19 - 4.17;p = 0.013, respectively) (**Figure 5, Supplemental Table 4**). The prevalence of 30% and 50% eGFR discrepancies by SMI quartiles by sex is shown in **Figure 5**.

We evaluated the continuous relationship between sarcopenia and the eGFR differences (eGFR_DIFF_) defined by subtracting eGFR_CRE_ from eGFR_CYS_ (negative values mean that eGFR_CYS_ is lower than eGFR_CRE_). The mean eGFR_DIFF_ was -20.9 ± 25.1 mL/min per 1.73m^2^ in patients with sarcopenia compared to -8.6 ± 22.4mL/min per 1.73m^2^ in patients without sarcopenia (p < 0.001). A spline model showing the continuous relationship between SMI and eGFR_DIFF_ demonstrated that as SMI decreases, the difference between eGFR_CRE_ and eGFR_CYS_ widened considerably (**Figure 6**). Heatmaps visualizing the relationship between eGFR_DIFF_ by SMI quartile and quartiles of total adiposity index, VAI, and SAI are shown in **Supplemental Figure 4**.

**Figure 6.**
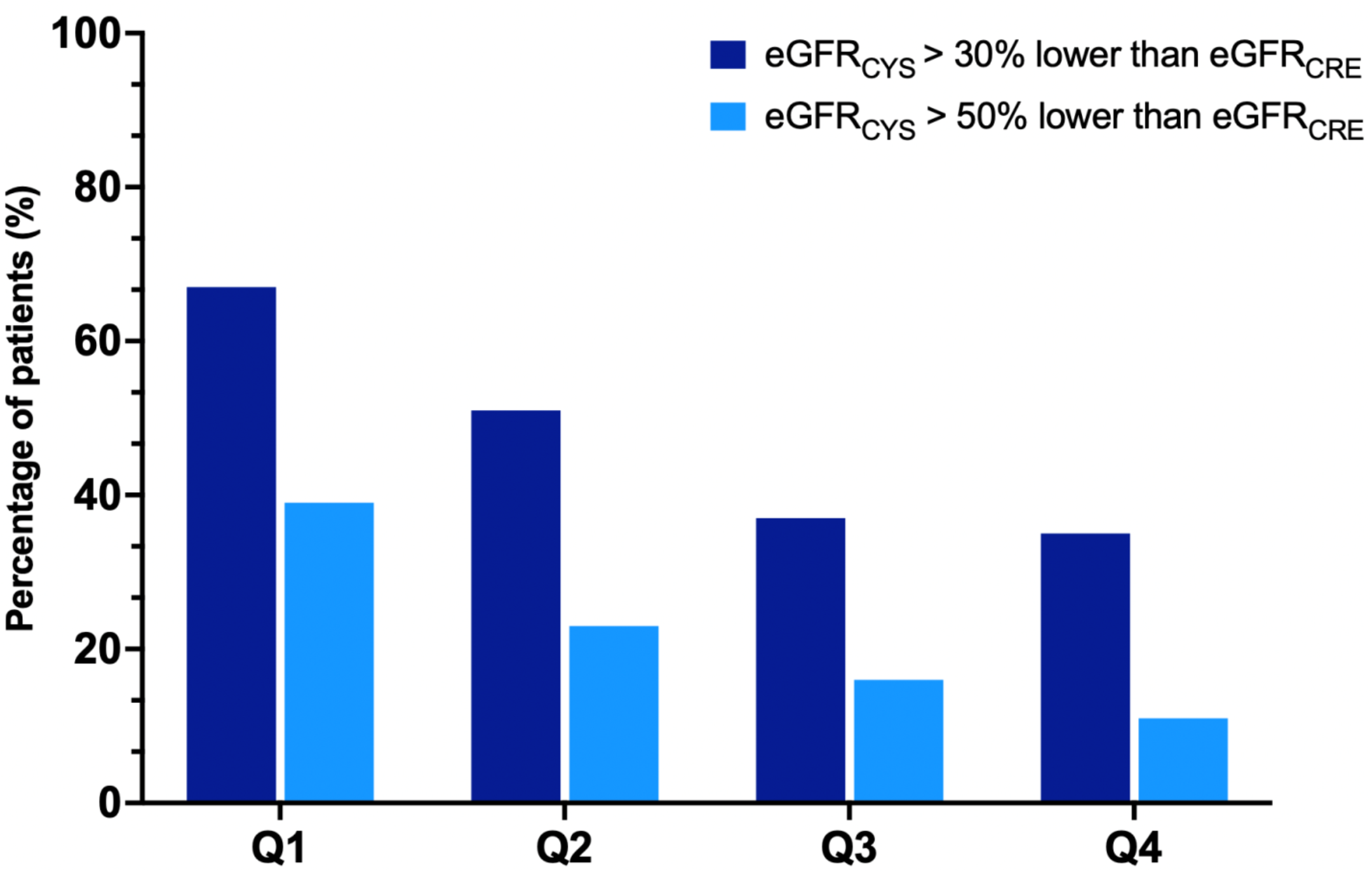
Rates of 30% and 50% eGFR discrepancies by skeletal muscle index quartile. Rates of >30% and >50% eGFR discrepancies by quartiles of skeletal muscle index. Lower SMI values (Q1) indicate a greater degree of sarcopenia. Abbreviations; eGFR = estimated glomerular filtration rate, SMI = skeletal muscle index, Q1 = first quartile, Q2 = second quartile, Q3 = third quartile, Q4 = 4^th^ quartile.

**Fig 7.**
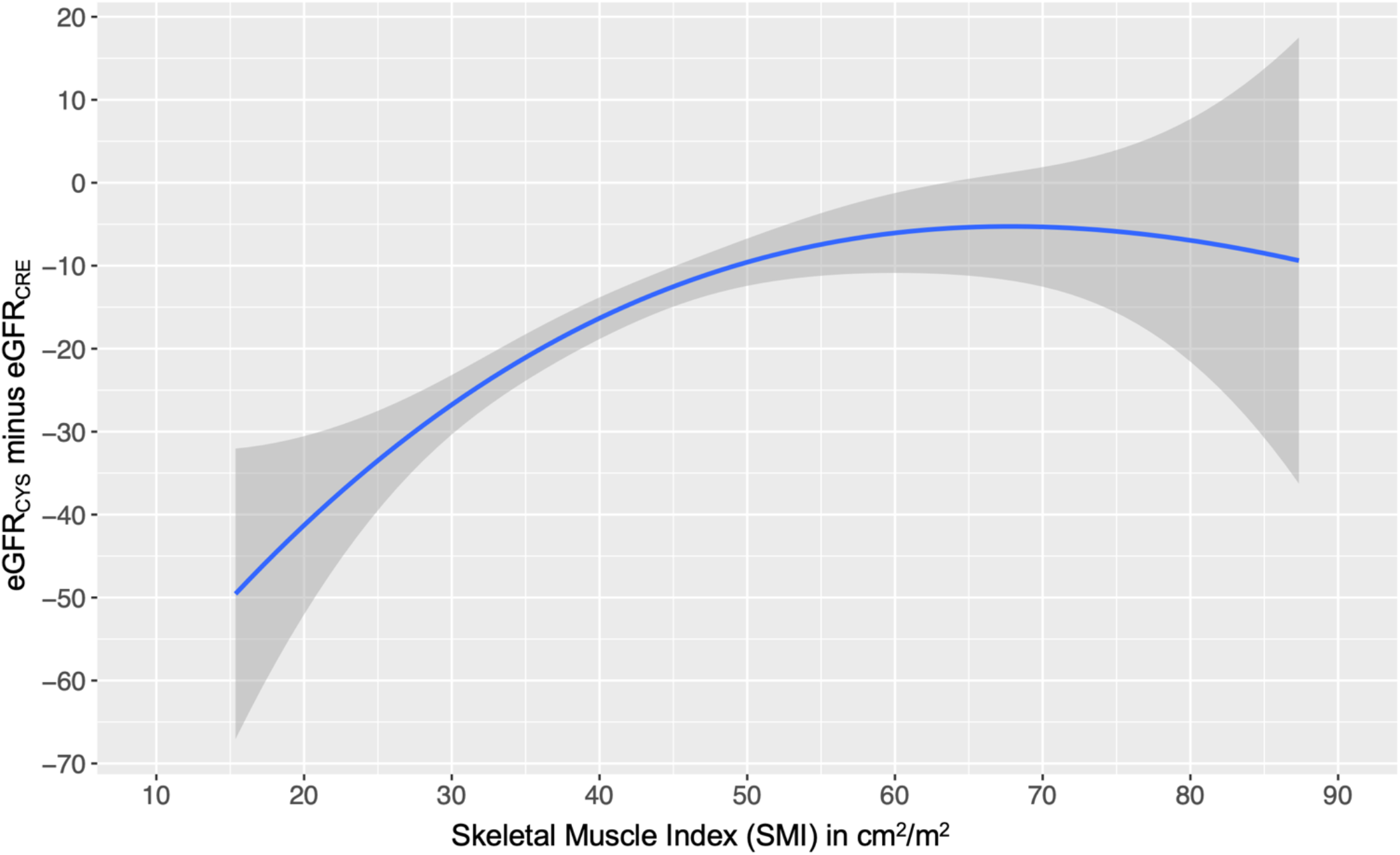
Spline model predicting the relationship between skeletal muscle index and eGFR difference (eGFR_CYS_ minus eGFR_CRE_) Unadjusted spline regression was performed evaluating the relationship between SMI and eGFR_DIFF_ (eGFR_CYS_ minus eGFR_CRE_), using B-Spline basis for cubic polynomial splines with 3 degrees of freedom using the 10^th^ to 90^th^ SMI percentile for the cohort. Lower SMI values indicate a greater degree of sarcopenia. Abbreviations: eGFR = estimated glomerular filtration rate, SMI = skeletal muscle index.

### Sensitivity analyses

Exclusion of patients with AKI, inpatient CT scans only, or liquid tumors did not meaningfully alter the relationship between SMI quartiles and eGFR discrepancy (**Supplemental Figure 5A, 5B and C**, respectively).

## Discussion

Our study demonstrates that imaging-defined sarcopenia and high adiposity were each independent risk factors for eGFR discrepancy in adult patients with a history of cancer. Moreover, as sarcopenia became more severe, the difference between eGFR_CRE_ and eGFR_CYS_ grew considerably larger (**Figure 6**).

Accurate assessment of eGFR is important, yet challenging in patients with cancer. Patients with cancer are commonly exposed to antineoplastic therapies or supportive medications (antibiotics, anticonvulsants, anticoagulants) that require dose adjustments based on kidney function; therefore, large discrepancies between eGFR_CRE_ and eGFR_CYS_ pose significant challenges in clinical management and may be associated with adverse clinical outcomes.^34^ In our study, the prevalence of eGFR discrepancies was high, emphasizing the importance of this knowledge gap. Large differences between eGFR_CRE_ and eGFR_CYS_ are associated with adverse clinical outcomes in several populations: a study of 9,092 hypertensive patients who participated in the Systolic Blood Pressure Intervention Trial (SPRINT) found that a more negative eGFR_DIFF_ (defined as eGFR_CYS_ - eGFR_CRE_) was associated with frailty, falls, hospitalizations, and death.^38^ Another study examining 4,512 patients with chronic kidney disease showed that having a large eGFR_DIFF_ was associated with an increased risk of hospitalization for new-onset heart failure.^39^ Of note, the rate of large eGFR discrepancies in our cohort was significantly higher than prior studies conducted in populations without cancer (**Supplemental Figure 3B**), suggesting that high prevalence of sarcopenia in patients with cancer could be an important risk factor for discrepancies between eGFR_CRE_ and eGFR_CYS_.^38,39^ A recent study in a cohort of 327 patients in China with resectable gastric cancer showed an association between lower serum creatine/cystatin C ratio and the presence of imaging-defined sarcopenia.^40^ We included a large cohort of patients with various cancer types and stages, and found a relationship between CT-defined sarcopenia and high adiposity with eGFR discrepancy.

Several non-GFR determinants for both cystatin C and SCr have been suggested in prior studies: sarcopenia is associated with lower SCr and can lead to overestimation of kidney function; while obesity is associated with higher cystatin C level and can lead to underestimation of kidney function by eGFR_CYS_.^8,11,41-43^ However, most prior studies utilized clinical and anthropomorphic measurements (BMI, muscle strength, skin fold thickness, etc.) as an assessment of sarcopenia and obesity, which may not as accurately reflect body composition of muscle and adipose tissue. Body composition analysis using CT scans is a more sensitive and specific way of assessing the quantity and distribution of skeletal muscle and adipose tissue than BMI.^12,16,17^ Deep learning models facilitate fully automated body composition analysis on routine abdominal CT scans, allowing for the potential for widespread clinical use in the future.^12,16,17,33^

Using independent sex-specific thresholds, we found that sarcopenia was common in our cohort: more than half (59%) met the imaging criteria for sarcopenia, in line with recent studies in patients with cancer, reporting a range of sarcopenia on imaging between 30 – 72%. ^44-47^ Sarcopenia is associated with decreased survival in patients with cancer and has been associated with increased treatment-related toxicity in patients receiving antineoplastic therapies.^48-51^ We hypothesize that medication overdose due to inaccurate estimation of GFR when relying on eGFR_CRE_ in patients with sarcopenia may cause dose-related chemotherapy toxicity, particularly with chemotherapies that are renally excreted,^48^ and may thereby contribute to the decreased survival in these patients.

Prior studies have suggested that obesity may increase cystatin C level independent of kidney function, which leads to underestimation of GFR using the cystatin C equation. However, this observation is largely based on obesity defined by BMI rather than detailed analysis of body composition.^8^ As cystatin C is secreted by nucleated cells and constitutively expressed in almost all organs,^52^ it is plausible to hypothesize that higher body cell mass, which correlates with higher BMI, could lead to higher cystatin C level independent of kidney function. A study in children showed that adjustment with body cell mass, estimated by bioimpedance, increased the accuracy of cystatin C-based GFR estimation.^53^ Other studies have suggested that increased cystatin C production in adipose tissue may be contributing to higher cystatin C level in obese individuals.^54^ In our study, we found that patients with high adiposity had higher odds of eGFR discrepancy. Future, large cohort studies are needed to validate our findings and to better understand the effect of subcutaneous and visceral adiposity on cystatin C. Finally, in our study, patients with liquid tumors had significantly higher rates of eGFR discrepancy than patients with solid tumors. This possibly reflects higher cancer cell burden in liquid tumors that associate with higher cystatin C level,^55^ suggesting the complicated relationship between serum biomarkers and individual disease states, and future studies are needed to best individualize GFR estimation in patients with cancer.

Our study has several limitations. First, cystatin C was ordered as part of clinical care which likely selected a population in whom clinicians questioned the accuracy of creatine eGFR estimation; therefore, the rate of eGFR discrepancy in our study is likely to be higher than in the general cancer population. Second, our study included a significant number of patients who were hospitalized or had unstable kidney function at the time of their CT scan, suggesting a significant fraction of patients included were experiencing acute illness. Third, we used a single measurement of SCr and cystatin C, which may not reflect a steady state at the time of measurement. Although these may limit the generalizability of our findings, these variables were adjusted for in the multivariable model, and sensitivity analyses excluding patients with inpatient CT scans and AKI did not meaningfully change the relationship between sarcopenia and eGFR discrepancy (**Supplemental Figure 4**). Fourth, a subset of patients either did not have a CT scan, or were excluded during quality assurance. Of note, automated segmentation was not possible in patients with soft tissue edema, which particularly affected the subcutaneous adipose tissue metric. Lastly, our study lacks the gold standard (measued GFR) given the retrospective design; prospective studies are needed to validate the accuracy of GFR estimating equations in consecutive patients with sarcopenia and high adiposity against measured GFR.

In conclusion, we demonstrate that sarcopenia is associated with higher risk of eGFR discrepancy; as SMI decreases, the discrepancy between eGFR_CRE_ and eGFR_CYS_ widens considerably. High adiposity is also associated with a higher risk of large eGFR discrepancies. Future studies are needed to improve and personalize the approach to GFR estimation in patients with cancer.

## Data Availability

De-identified data produced in the present study are available upon reasonable request to the corresponding author.

---

**Supplemental Table 1.** Reasons for body composition analysis failure. There were 48 patients whose scans were excluded from body composition analysis for reasons listed in the above table.

| **Reason** | **N = 48** |
| --- | --- |
| **i. Artifacts due to low dose technique, motion, or image reconstruction** | 23 |
| **ii. Soft tissue edema** | 9 |
| **iii. Small field of view** | 6 |
| **iv. Artifacts from surgical hardware, drains, or abdominal wall hernia** | 6 |
| **v. Artifacts from patient skin touching CT gantry** | 4 |

**Supplemental Table 2.** Cancer types by skeletal muscle index quartile (SMI), in alphabetical order. Cancer types by skeletal muscle index quartile (SMI). *Primary cancer type was determined by the type most frequently coded in the electronic health record.

| Cancer Type | Overall | 1<sup>st</sup> Quartile of SMI | 2<sup>nd</sup> Quartile of SMI | 3<sup>rd</sup> Quartile of SMI | 4<sup>th</sup> Quartile of SMI |
| --- | --- | --- | --- | --- | --- |
|  | N= 545 | N= 137 | N= 136 | N= 136 | N= 136 |
| **Brain** | 8 (1.5%) | 1 (0.7%) | 1 (0.7%) | 3 (2.2%) | 3 (2.2%) |
| **Breast** | 68 (12%) | 12 (8.8%) | 23 (17%) | 15 (11%) | 18 (13%) |
| **Gastrointestinal** | 59 (11%) | 15 (11%) | 15 (11%) | 15 (11%) | 14 (10%) |
| **Genitourinary Cancer** | 55 (10.1%) | 10 (7.3%) | 16 (11.8%) | 12 (8.8%) | 17 (12.5%) |
| **Gynecological** | 62 (11%) | 12 (8.8%) | 12 (8.8%) | 20 (15%) | 18 (13%) |
| **Head & Neck cancer** | 12 (2.2%) | 4 (2.9%) | 3 (2.2%) | 1 (0.7%) | 4 (2.9%) |
| **Leukemia** | 54 (9.9%) | 20 (15%) | 14 (10%) | 17 (12%) | 3 (2.2%) |
| **Lung cancer** | 51 (9.4%) | 17 (12%) | 13 (9.6%) | 11 (8.1%) | 10 (7.4%) |
| **Lymphoma** | 46 (8.4%) | 13 (9.5%) | 15 (11%) | 5 (3.7%) | 13 (9.6%) |
| **Melanoma** | 4 (0.7%) | 1 (0.7%) | 3 (2.2%) | 0 (0%) | 0 (0%) |
| **Non-melanomatous skin cancer** | 41 (7.5%) | 12 (8.8%) | 6 (4.4%) | 8 (5.9%) | 15 (11%) |
| **Renal cell carcinoma** | 39 (7.2%) | 9 (6.6%) | 5 (3.7%) | 17 (12%) | 8 (5.9%) |
| **Sarcoma** | 1 (0.2%) | 1 (0.7%) | 0 (0%) | 0 (0%) | 0 (0%) |
| **Thyroid cancer** | 8 (1.5%) | 2 (1.5%) | 4 (2.9%) | 0 (0%) | 2 (1.5%) |
| **Other** | 37 (6.8%) | 8 (5.8%) | 6 (4.4%) | 12 (8.8%) | 11 (8.1%) |

**Supplemental Table 3.** Comparison of baseline characteristics of patients with excluded vs. included scans. The baseline characteristics of the 48 patients whose scans were excluded from body composition analysis are compared to the included cohort. The reason for scan exclusion is shown in **Supplemental Table 1**.

| **Covariates** | **Excluded scans** | **Included scans** |
| --- | --- | --- |
|  | N = 48 | N = 545 |
| **Age, years** | 72 [62, 79] | 63 [55, 72] |
| **Male Sex** | 22 (45.8%) | 245 (45.0%) |
| **White Race** | 32 (66.7%) | 440 (80.7%) |
| **Body Mass Index** |  |  |
| **Normal (18.5 - 24.9)** | 13 (27.1%) | 190 (34.9%) |
| **Underweight (<18.5)** | 2 (4.2%) | 30 (5.5%) |
| **Overweight (25 - 29.9)** | 27 (56.2%) | 164 (30.1%) |
| **Obese (≥30)** | 6 (12.5%) | 161 (29.5%) |
| **Baseline eGFR<sub>CRE-CYS</sub>** | 43.9 [23.6, 69.8] | 46.3 [30.7, 75.3] |
| **Advanced/Active treatment** | 25(52.1%) | 338(62%) |
| **Early-stage/Remission** | 23 (47.9%) | 207 (38.0%) |
| **Comorbidities** |  |  |
| **Hypertension** | 43 (89.6%) | 408 (74.9%) |
| **Coronary Artery Disease** | 36 (75.0%) | 279 (51.2%) |
| **Diabetes Mellitus** | 34 (70.8%) | 299 (54.9%) |
| **Cirrhosis** | 4 (8.3%) | 43 (7.9%) |
| **Human Immunodeficiency Virus** | 1 (2.1%) | 20 (3.7%) |
| **Smoking** | 19 (39.6%) | 235 (43.1%) |
| **Malnutrition** | 10 (20.8%) | 68 (12.5%) |
| **Labs** |  |  |
| **Serum Albumin (g/dL)** |  |  |
| **<3.0** | 35 (72.9%) | 168 (30.8%) |
| **3.0-3.9** | 7 (14.6%) | 184 (33.8%) |
| **≥4.0** | 6 (12.5%) | 193 (35.4%) |
| **Serum Creatinine (mg/dL)** | 1.06 [0.72, 1.68] | 1.24 [0.85, 1.73] |
| **Serum Cystatin C (mg/L)** | 1.96 [1.12, 3.12] | 1.63 [1.12, 2.33] |
| **Hemoglobin (g/dL)** |  |  |
| **<10.0** | 34 (70.8%) | 274 (50.3%) |
| **10.0 - 11.9** | 9 (18.8%) | 131 (24.0%) |
| **≥12.0** | 5 (10.4%) | 140 (25.7%) |

**Supplemental Table 4.** Predictors of eGFR_CYS_ more than 50% lower than eGFR_CRE_. Baseline eGFR was defined using the race-free CKD Epi 2021 combined cystatin C and creatinine equation. Abbreviations: CI = confidence interval, REF = reference, ACEi/ARB = angiotensin converting enzyme inhibitor or angiotensin II receptor blockade, HIV = human immunodeficiency virus.

| Covariates | eGFR<sub>CYS</sub> >50% lower than eGFR<sub>CRE</sub><br>(N=122 vs. N=423 [reference]) |  |  |  |  |  |
| --- | --- | --- | --- | --- | --- | --- |
|  | Univariable |  |  | Multivariable |  |  |
|  | OR | 95% CI | p-value | Adj OR | 95% CI | p-value |
| Age (per 10 years) | 1.04 | 0.90,1.20 | 0.619 | 0.82 | 0.66, 1.01 | 0.057 |
| Male Sex | 1.83 | 1.22,2.75 | 0.004 | 1.12 | 0.63, 2.00 | 0.696 |
| Baseline eGFR | 0.98 | 0.97,0.98 | <0.001 | 0.98 | 0.97, 0.99 | <0.001 |
| Sarcopenia | 3.28 | 2.04,5.27 | <0.001 | 2.34 | 1.21, 4.51 | 0.012 |
| Highest adiposity quartile | 1.28 | 0.80,2.02 | 0.300 | 2.23 | 1.19, 4.17 | 0.013 |
| Liquid vs. solid tumor | 3.50 | 2.19,5.57 | <0.001 | 1.69 | 0.89, 3.22 | 0.110 |
| Acute kidney injury | 2.30 | 1.53,3.47 | <0.001 | 0.71 | 0.41, 1.24 | 0.226 |
| Inpatient vs. Outpatient CT scan | 4.56 | 2.98,6.97 | <0.001 | 2.04 | 1.19, 3.51 | 0.010 |
| Smoking | 1.56 | 1.04,2.34 | 0.0320 | 1.34 | 0.79, 2.28 | 0.284 |
| **Comorbidities** |  |  |  |  |  |  |
| Hypertension | 2.10 | 1.23,3.58 | 0.007 | 0.72 | 0.32, 1.64 | 0.438 |
| Coronary Artery Disease | 3.31 | 2.12,5.16 | <0.001 | 1.80 | 1.00, 3.24 | 0.049 |
| Diabetes Mellitus | 5.47 | 3.29,9.09 | <0.001 | 2.17 | 1.14, 4.11 | 0.018 |
| Cirrhosis | 4.21 | 2.22,7.97 | <0.001 | 3.19 | 1.43, 7.11 | 0.005 |
| HIV | 2.40 | 0.96,6.03 | 0.062 |  |  |  |
| Malnutrition | 1.53 | 0.87,2.70 | 0.140 |  |  |  |
| Thyroid disease | 1.30 | 0.83,2.03 | 0.245 |  |  |  |
| **Medication Use** |  |  |  |  |  |  |
| Corticosteroids | 2.98 | 1.97,4.51 | <0.001 | 1.04 | 0.61, 1.77 | 0.897 |
| **Labs** |  |  |  |  |  |  |
| Albumin (g/dL) |  |  |  |  |  |  |
| ≥4.0 | REF | — |  | — | — |  |
| 3.0-3.9 | 8.17 | 3.36, 19.9 | <0.001 | 3.92 | 1.34, 11.4 | 0.013 |
| <3.0 | 27.1 | 11.4, 64.7 | <0.001 | 11.9 | 4.03, 35.0 | <0.001 |
| Hemoglobin (g/dL) |  |  |  |  |  |  |
| ≥12.0 | REF | — |  | — | — |  |
| 10.0 - 11.9 | 2.13 | 0.87, 5.22 | 0.097 | 0.72 | 0.24,2.18 | 0.565 |
| <10.0 | 9.33 | 4.38, 19.9 | <0.001 | 1.47 | 0.54, 4.03 | 0.455 |

**Supplemental Figure 1.**
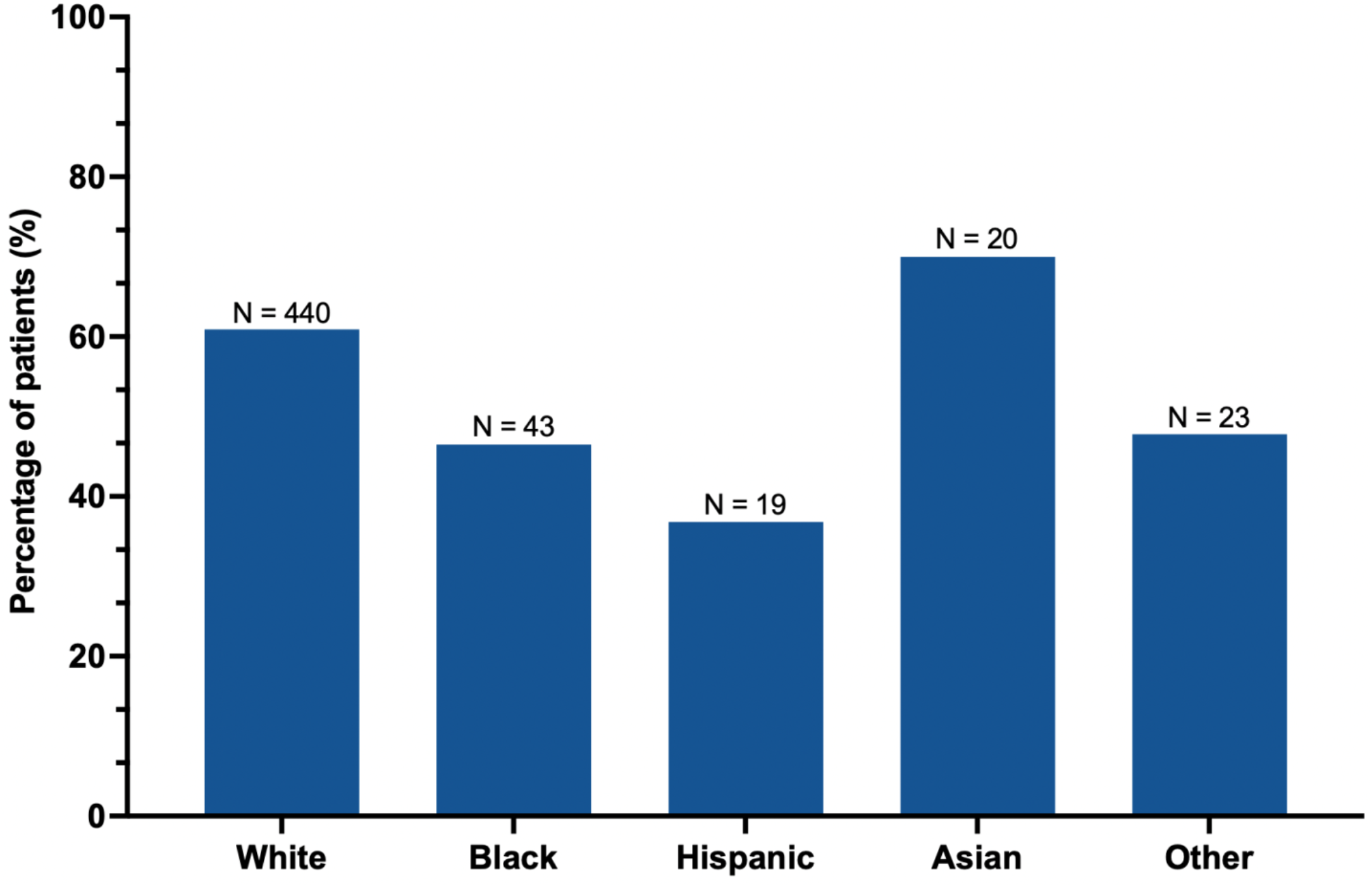
Sarcopenia by race/ethnicity. Sarcopenia by self-reported race/ethnicity

**Supplemental Figure 2.**
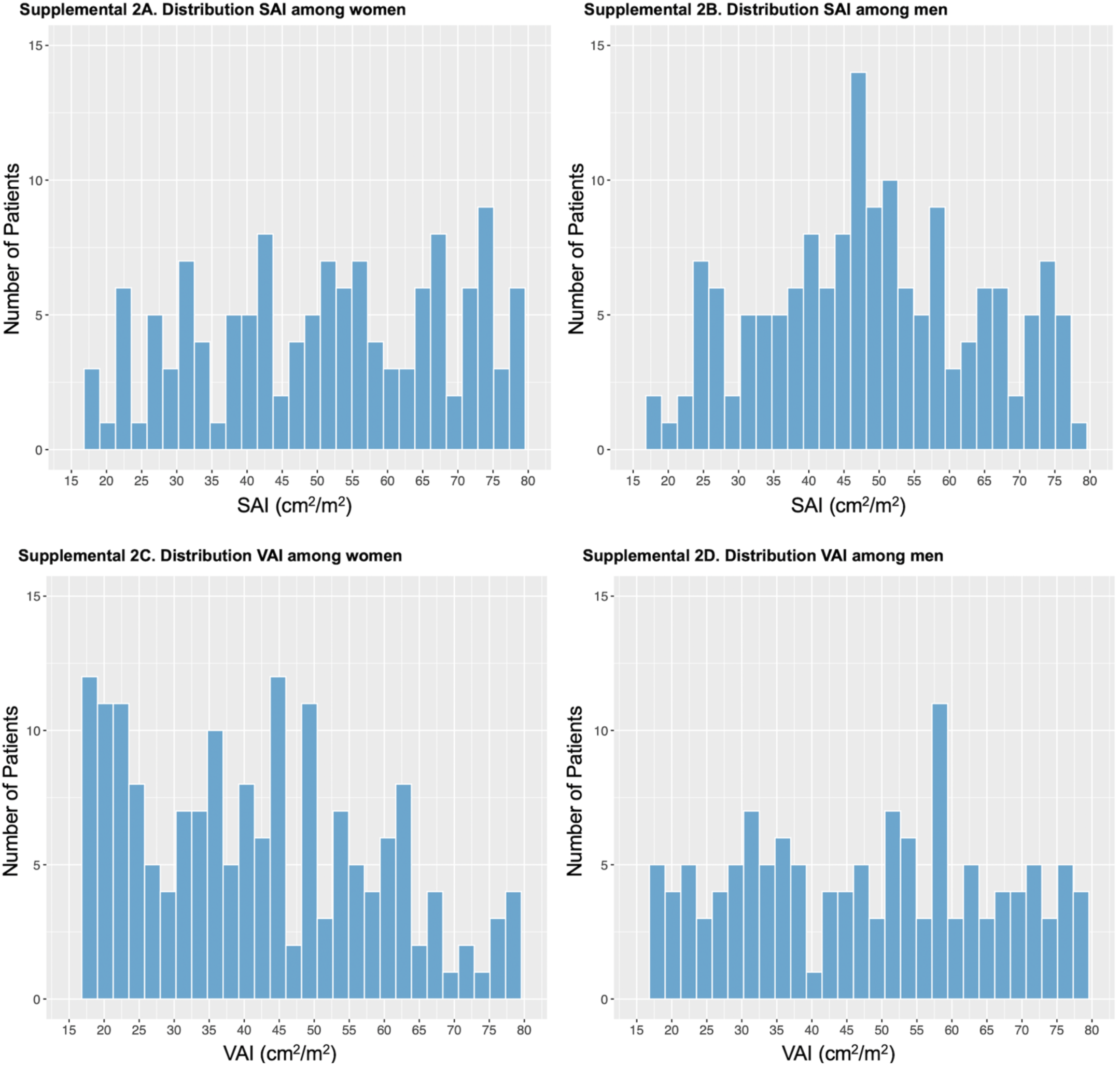
Sex-stratified histograms of subcutaneous and visceral adiposity indices. Subcutaneous and visceral adiposity stratified by sex. Abbreviations; SAI = subcutaneous adiposity index, VAI = visceral adiposity index.

**Supplemental Figure 3.**
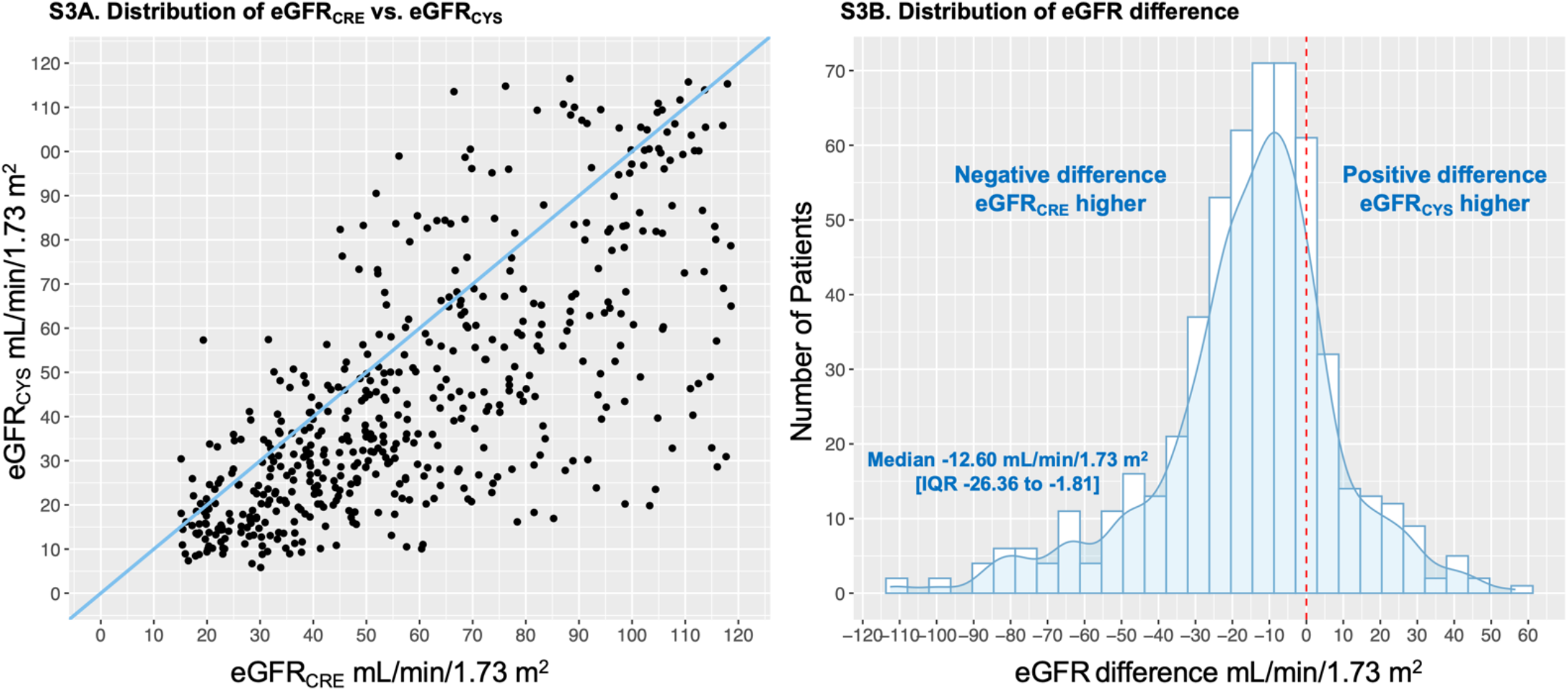
Scatter plot of creatinine-based and cystatin C-based eGFR, and distribution of eGFR difference. Figure S3A. Scatterplot showing distribution of eGFR_CRE_ and eGFR_CYS_ among study patients; the blue line is the line of equality. Figure S3B. A histogram and superimposed density curve showing the distribution of eGFR difference defined by eGFR_CYS_ minus eGFR_CRE_. The red dotted line signifies equivalence between eGFR_CRE_ and eGFR_CYS_. Abbreviations: eGFR = estimated glomerular filtration rate, IQR = interquartile range

**Supplemental Figure 4.**
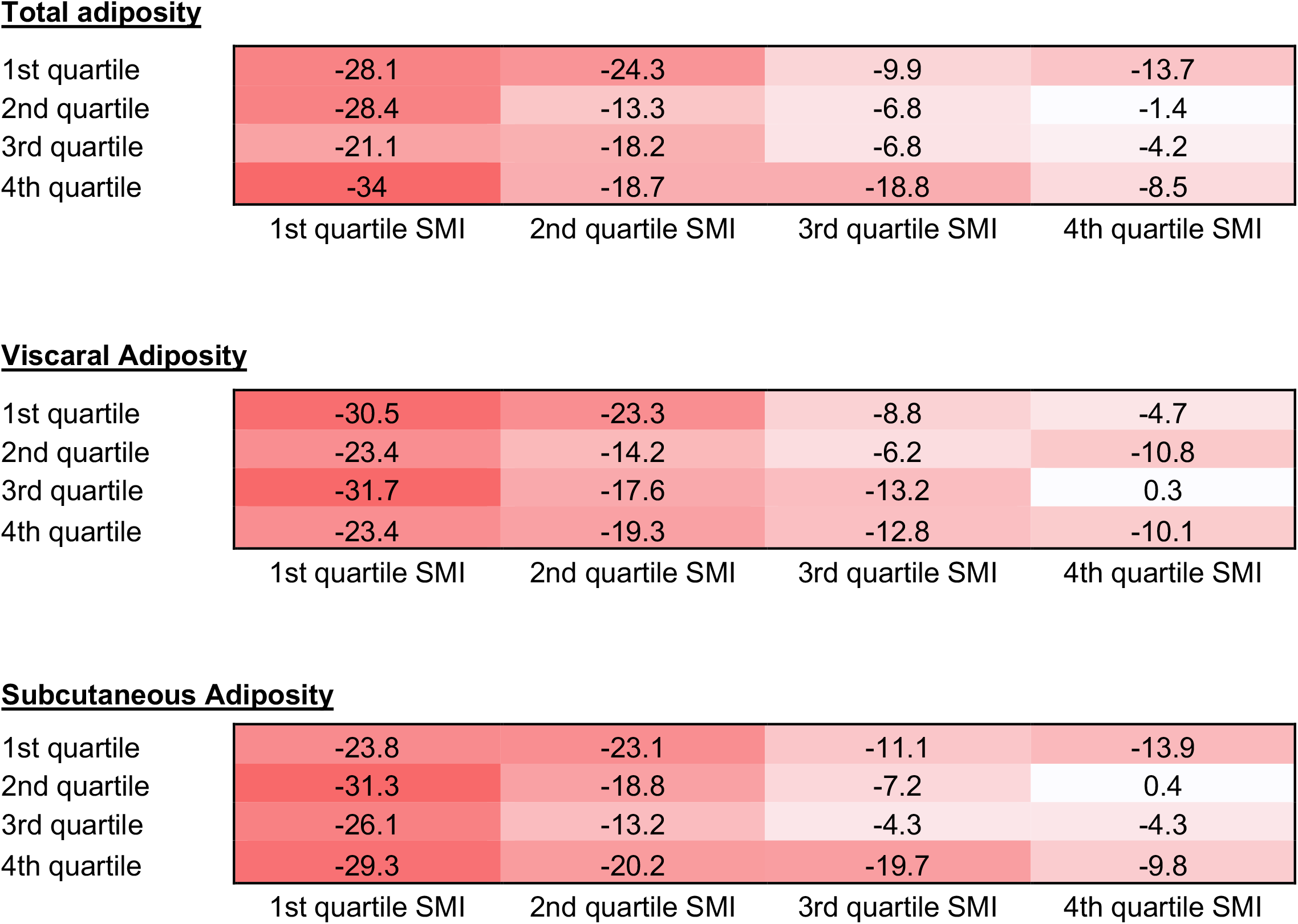
Heatmaps showing eGFR difference by quartile of total, visceral, and subcutaneous adiposity, and skeletal mass index. Values in each cell are the mean eGFR difference (defined by eGFR_CYS_ minus eGFR_CRE_) in mL/min/1.73m^2^ by quartile. Abbreviations: SMI = skeletal mass index.

**Supplemental Figure 5.**
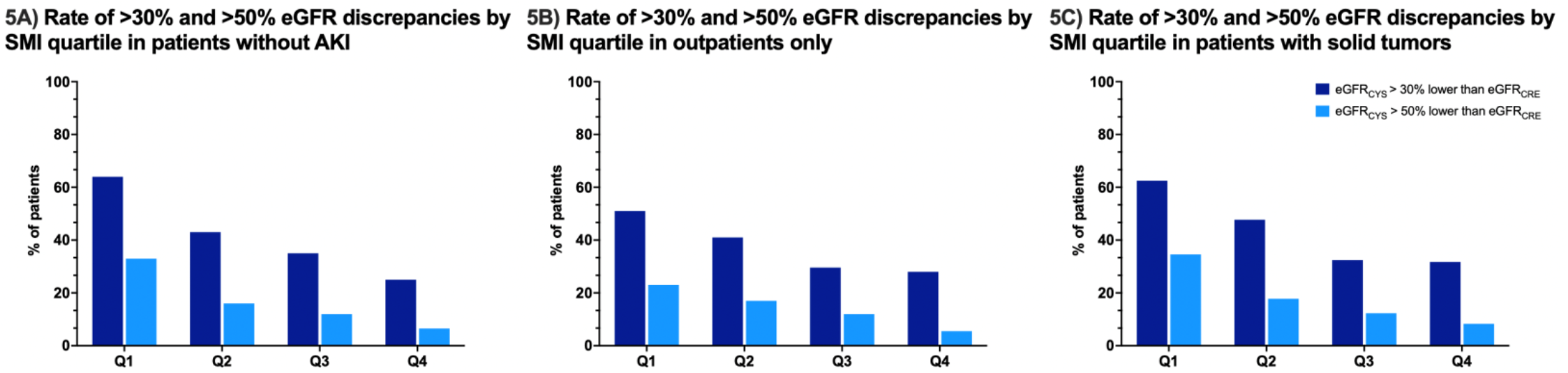
Sensitivity analyses. 5A. Sensitivity analysis in patients without acute kidney injury at time of creatinine/cystatin C check, N=319. 5B. patients with outpatient scans, N = 375 and 5C. patients with solid tumors only N = 445 (5C). Abbreviations: eGFR = estimated glomerular filtration rate, SMI = skeletal muscle index, AKI = acute kidney injury, Q1 = first quartile, Q2 = second quartile, Q3 = third quartile, Q4 = 4^th^ quartile.

## References

1. Levey AS, Bosch JP, Lewis JB, Greene T, Rogers N, Roth D. A more accurate method to estimate glomerular filtration rate from serum creatinine: a new prediction equation. Modification of Diet in Renal Disease Study Group. Ann Intern Med. Mar 16 1999;130(6):461–70. doi:10.7326/0003-4819-130-6-199903160-00002

2. Levey AS, Coresh J, Greene T, et al. Using standardized serum creatinine values in the modification of diet in renal disease study equation for estimating glomerular filtration rate. Ann Intern Med. Aug 15 2006;145(4):247–54. doi:10.7326/0003-4819-145-4-200608150-00004

3. Cockcroft DW, Gault MH. Prediction of creatinine clearance from serum creatinine. Nephron. 1976;16(1):31–41. doi:10.1159/000180580

4. Inker LA, Eneanya ND, Coresh J, et al. New Creatinine- and Cystatin C-Based Equations to Estimate GFR without Race. N Engl J Med. Nov 4 2021;385(19):1737–1749. doi:10.1056/NEJMoa2102953

5. Shafi T, Zhu X, Lirette ST, et al. Quantifying Individual-Level Inaccuracy in Glomerular Filtration Rate Estimation : A Cross-Sectional Study. Ann Intern Med. Aug 2022;175(8):1073–1082. doi:10.7326/M22-0610

6. Stevens LA, Schmid CH, Greene T, et al. Factors other than glomerular filtration rate affect serum cystatin C levels. Kidney Int. Mar 2009;75(6):652–60. doi:10.1038/ki.2008.638

7. Inker LA, Schmid CH, Tighiouart H, et al. Estimating glomerular filtration rate from serum creatinine and cystatin C. N Engl J Med. Jul 5 2012;367(1):20–9. doi:10.1056/NEJMoa1114248

8. Knight EL, Verhave JC, Spiegelman D, et al. Factors influencing serum cystatin C levels other than renal function and the impact on renal function measurement. Kidney Int. Apr 2004;65(4):1416–21. doi:10.1111/j.1523-1755.2004.00517.x

9. Bruggeman AR, Kamal AH, LeBlanc TW, Ma JD, Baracos VE, Roeland EJ. Cancer Cachexia: Beyond Weight Loss. Journal of Oncology Practice. 2016;12(11):1163–1171. doi:10.1200/jop.2016.016832

10. Zamboni M, Mazzali G, Fantin F, Rossi A, Di Francesco V. Sarcopenic obesity: a new category of obesity in the elderly. Nutr Metab Cardiovasc Dis. Jun 2008;18(5):388–95. doi:10.1016/j.numecd.2007.10.002

11. Baracos VE, Arribas L. Sarcopenic obesity: hidden muscle wasting and its impact for survival and complications of cancer therapy. Ann Oncol. Feb 1 2018;29(Suppl_2):ii1–ii9. doi:10.1093/annonc/mdx810

12. Prado CM, Lieffers JR, McCargar LJ, et al. Prevalence and clinical implications of sarcopenic obesity in patients with solid tumours of the respiratory and gastrointestinal tracts: a population-based study. Lancet Oncol. Jul 2008;9(7):629–35. doi:10.1016/S1470-2045(08)70153-0

13. Martin L, Birdsell L, Macdonald N, et al. Cancer cachexia in the age of obesity: skeletal muscle depletion is a powerful prognostic factor, independent of body mass index. J Clin Oncol. Apr 20 2013;31(12):1539–47. doi:10.1200/jco.2012.45.2722

14. Ross R, Janssen I. Computed Tomography and Magnetic Resonance Imaging. In: Heymsfield SB, Lohman TG, Wang Z, Going SB, eds. Human Body Composition. 2 ed. Human Kinetics; 2005:89–108.

15. Loan MDV. Human Body Composition 2nd ed, edited by SB Heymsfield, TG Lohman, ZM Wang, and SB Going, 2005, 536 pages, hardcover, $89. Human Kinetics, Champaign, IL. The American Journal of Clinical Nutrition. 2005;82:1361–1361.

16. Shen W, Punyanitya M, Wang Z, et al. Visceral adipose tissue: relations between single-slice areas and total volume. The American journal of clinical nutrition. 2004;80(2):271–278. doi:10.1093/ajcn/80.2.271

17. Shen W, Punyanitya M, Wang Z, et al. Total body skeletal muscle and adipose tissue volumes: estimation from a single abdominal cross-sectional image. J Appl Physiol (1985). Dec 2004;97(6):2333–8. doi:10.1152/japplphysiol.00744.2004

18. Dalal S, Hui D, Bidaut L, et al. Relationships among body mass index, longitudinal body composition alterations, and survival in patients with locally advanced pancreatic cancer receiving chemoradiation: a pilot study. J Pain Symptom Manage. Aug 2012;44(2):181–91. doi:10.1016/j.jpainsymman.2011.09.010

19. Malietzis G, Currie AC, Athanasiou T, et al. Influence of body composition profile on outcomes following colorectal cancer surgery. Br J Surg. Apr 2016;103(5):572–80. doi:10.1002/bjs.10075

20. Rier HN, Jager A, Sleijfer S, van Rosmalen J, Kock Mcjm, Levin M-D. Low muscle attenuation is a prognostic factor for survival in metastatic breast cancer patients treated with first line palliative chemotherapy. The Breast. 2017/02/01/ 2017;31:9–15. doi:https://doi.org/10.1016/j.breast.2016.10.014

21. Cushen SJ, Power DG, Murphy KP, et al. Impact of body composition parameters on clinical outcomes in patients with metastatic castrate-resistant prostate cancer treated with docetaxel. Clin Nutr ESPEN. Jun 2016;13:e39–e45. doi:10.1016/j.clnesp.2016.04.001

22. Sandini M, Patiňo M, Ferrone CR, et al. Association Between Changes in Body Composition and Neoadjuvant Treatment for Pancreatic Cancer. JAMA Surgery. 2018;153(9):809–815. doi:10.1001/jamasurg.2018.0979

23. van Seventer EE, Fintelmann FJ, Roeland EJ, Nipp RD. Leveraging the Potential Synergy Between Patient-Reported Outcomes and Body Composition Analysis in Patients with Cancer. Oncologist. Apr 2020;25(4):271–273. doi:10.1634/theoncologist.2019-0813

24. DeFilipp Z, Troschel FM, Qualls DA, et al. Evolution of Body Composition Following Autologous and Allogeneic Hematopoietic Cell Transplantation: Incidence of Sarcopenia and Association with Clinical Outcomes. Biol Blood Marrow Transplant. Aug 2018;24(8):1741–1747. doi:10.1016/j.bbmt.2018.02.016

25. Best TD, Roeland EJ, Horick NK, et al. Muscle Loss Is Associated with Overall Survival in Patients with Metastatic Colorectal Cancer Independent of Tumor Mutational Status and Weight Loss. Oncologist. Jun 2021;26(6):e963–e970. doi:10.1002/onco.13774

26. Chute DF, Zhao S, Strohbehn IA, et al. Incidence and Predictors of CKD and Estimated GFR Decline in Patients Receiving Immune Checkpoint Inhibitors. Am J Kidney Dis. Jan 2022;79(1):134–137. doi:10.1053/j.ajkd.2021.05.012

27. Seethapathy H, Zhao S, Chute DF, et al. The Incidence, Causes, and Risk Factors of Acute Kidney Injury in Patients Receiving Immune Checkpoint Inhibitors. Clin J Am Soc Nephrol. Dec 6 2019;14(12):1692–1700. doi:10.2215/CJN.00990119

28. Inker LA, Eckfeldt J, Levey AS, et al. Expressing the CKD-EPI (Chronic Kidney Disease Epidemiology Collaboration) cystatin C equations for estimating GFR with standardized serum cystatin C values. Am J Kidney Dis. Oct 2011;58(4):682–4. doi:10.1053/j.ajkd.2011.05.019

29. Stevens LA, Coresh J, Schmid CH, et al. Estimating GFR using serum cystatin C alone and in combination with serum creatinine: a pooled analysis of 3,418 individuals with CKD. Am J Kidney Dis. Mar 2008;51(3):395–406. doi:10.1053/j.ajkd.2007.11.018

30. van Seventer E, Marquardt JP, Troschel AS, et al. Associations of Skeletal Muscle With Symptom Burden and Clinical Outcomes in Hospitalized Patients With Advanced Cancer. J Natl Compr Canc Netw. Jan 29 2021;19(3):319–327. doi:10.6004/jnccn.2020.7618

31. Bridge CP, Rosenthal M, Wright B, et al. Fully-Automated Analysis of Body Composition from CT in Cancer Patients Using Convolutional Neural Networks. Springer International Publishing; 2018:204–213.

32. Bridge CP, Rosenthal M, Wright B, et al. Fully-automated analysis of body composition from CT in cancer patients using convolutional neural networks. OR 20 Context-aware operating theaters, computer assisted robotic endoscopy, clinical image-based procedures, and skin image analysis. Springer; 2018:204–213.

33. Magudia K, Bridge CP, Bay CP, et al. Population-Scale CT-based Body Composition Analysis of a Large Outpatient Population Using Deep Learning to Derive Age-, Sex-, and Race-specific Reference Curves. Radiology. 2021;298(2):319–329. doi:10.1148/radiol.2020201640

34. Hanna PE, Wang Q, Strohbehn I, et al. Medication-related adverse events in patients with cancer and discrepancies in cystatin C-versus creatinine-based eGFR. medRxiv. 2023:2023.01.18.23284656. doi:10.1101/2023.01.18.23284656

35. Costa Esvt, Gil LA, Jr., Inker LA, et al. A prospective cross-sectional study estimated glomerular filtration rate from creatinine and cystatin C in adults with solid tumors. Kidney Int. Mar 2022;101(3):607–614. doi:10.1016/j.kint.2021.12.010

36. Delgado C, Baweja M, Crews DC, et al. A Unifying Approach for GFR Estimation: Recommendations of the NKF-ASN Task Force on Reassessing the Inclusion of Race in Diagnosing Kidney Disease. J Am Soc Nephrol. Sep 23 2021;32(12):2994–3015. doi:10.1681/asn.2021070988

37. Rubin D. Multiple Imputation for Nonresponse in Surveys. New York, NY: JohnWiley & Sons. Inc; 1987.

38. Potok OA, Ix JH, Shlipak MG, et al. The Difference Between Cystatin C- and Creatinine-Based Estimated GFR and Associations With Frailty and Adverse Outcomes: A Cohort Analysis of the Systolic Blood Pressure Intervention Trial (SPRINT). Am J Kidney Dis. Dec 2020;76(6):765–774. doi:10.1053/j.ajkd.2020.05.017

39. Chen DC, Shlipak MG, Scherzer R, et al. Association of Intra-individual Differences in Estimated GFR by Creatinine Versus Cystatin C With Incident Heart Failure. Am J Kidney Dis. Dec 2022;80(6):762–772 e1. doi:10.1053/j.ajkd.2022.05.011

40. Sun J, Yang H, Cai W, et al. Serum creatinine/cystatin C ratio as a surrogate marker for sarcopenia in patients with gastric cancer. BMC Gastroenterology. 2022/01/19 2022;22(1):26. doi:10.1186/s12876-022-02093-4

41. Hudson JQ, Nolin TD. Pragmatic Use of Kidney Function Estimates for Drug Dosing: The Tide Is Turning. Adv Chronic Kidney Dis. Jan 2018;25(1):14–20. doi:10.1053/j.ackd.2017.10.003

42. Yoshida S, Suda G, Ohara M, et al. Frequency and Characteristics of Overestimated Renal Function in Japanese Patients with Chronic Liver Disease and Its Relation to Sarcopenia. Nutrients. Jul 14 2021;13(7)doi:10.3390/nu13072415

43. Williams GR, Dunne RF, Giri S, Shachar SS, Caan BJ. Sarcopenia in the Older Adult With Cancer. J Clin Oncol. Jul 1 2021;39(19):2068–2078. doi:10.1200/jco.21.00102

44. Suzuki Y, Okamoto T, Fujishita T, et al. Clinical implications of sarcopenia in patients undergoing complete resection for early non-small cell lung cancer. Lung Cancer. Nov 2016;101:92–97. doi:10.1016/j.lungcan.2016.08.007

45. Choi H, Park YS, Na KJ, et al. Association of Adipopenia at Preoperative PET/CT with Mortality in Stage I Non-Small Cell Lung Cancer. Radiology. Dec 2021;301(3):645–653. doi:10.1148/radiol.2021210576

46. Deluche E, Lachatre D, Di Palma M, et al. Is sarcopenia a missed factor in the management of patients with metastatic breast cancer? The Breast. 2022/02/01/ 2022;61:84–90. doi:https://doi.org/10.1016/j.breast.2021.12.014

47. Catikkas NM, Bahat Z, Oren MM, Bahat G. Older cancer patients receiving radiotherapy: a systematic review for the role of sarcopenia in treatment outcomes. Aging Clinical and Experimental Research. 2022/08/01 2022;34(8):1747–1759. doi:10.1007/s40520-022-02085-0

48. Prado CM, Baracos VE, McCargar LJ, et al. Sarcopenia as a determinant of chemotherapy toxicity and time to tumor progression in metastatic breast cancer patients receiving capecitabine treatment. Clin Cancer Res. Apr 15 2009;15(8):2920–6. doi:10.1158/1078-0432.CCR-08-2242

49. Prado CM, Baracos VE, McCargar LJ, et al. Body composition as an independent determinant of 5-fluorouracil-based chemotherapy toxicity. Clin Cancer Res. Jun 1 2007;13(11):3264–8. doi:10.1158/1078-0432.CCR-06-3067

50. Mir O, Coriat R, Blanchet B, et al. Sarcopenia predicts early dose-limiting toxicities and pharmacokinetics of sorafenib in patients with hepatocellular carcinoma. PLoS One. 2012;7(5):e37563. doi:10.1371/journal.pone.0037563

51. Antoun S, Baracos VE, Birdsell L, Escudier B, Sawyer MB. Low body mass index and sarcopenia associated with dose-limiting toxicity of sorafenib in patients with renal cell carcinoma. Ann Oncol. Aug 2010;21(8):1594–1598. doi:10.1093/annonc/mdp605

52. Abrahamson M, Olafsson I, Palsdottir A, et al. Structure and expression of the human cystatin C gene. Biochem J. Jun 1 1990;268(2):287–94. doi:10.1042/bj2680287

53. Andersen TB, Jødal L, Boegsted M, et al. GFR Prediction From Cystatin C and Creatinine in Children: Effect of Including Body Cell Mass. American Journal of Kidney Diseases. 2012/01/01/ 2012;59(1):50–57. doi:https://doi.org/10.1053/j.ajkd.2011.09.013

54. Naour N, Fellahi S, Renucci JF, et al. Potential contribution of adipose tissue to elevated serum cystatin C in human obesity. Obesity (Silver Spring). Dec 2009;17(12):2121–6. doi:10.1038/oby.2009.96

55. Hammouda NE, Salah El-Din Ma, El-Shishtawy MM, El-Gayar AM. Serum Cystatin C as a Biomarker in Diffuse Large B-Cell Lymphoma. Sci Pharm. Mar 8 2017;85(1)doi:10.3390/scipharm85010009

